# Impact of Congestive Heart Failure and Role of Cardiac Biomarkers in COVID-19 patients: A Systematic Review and Meta-Analysis

**DOI:** 10.1101/2020.07.06.20147421

**Authors:** Tarun Dalia, Shubham Lahan, Sagar Ranka, Prakash Acharya, Archana Gautam, Ioannis Mastoris, Andrew Sauer, Zubair Shah

## Abstract

**Importance:** Coronavirus disease 2019 (COVID-19) has been reported to cause worse outcomes in patients with underlying cardiovascular disease, especially in patients with acute cardiac injury, which is determined by elevated levels of high-sensitivity troponin. There is paucity of data on the impact of congestive heart failure (CHF) on outcomes in COVID-19 patients.

**Objective:** To evaluate the occurrence of acute cardiac injury and arrhythmias and to assess the impact of pre-existing CHF and hypertension (HTN) in COVID-19 patients.

**Data Sources:** We conducted a literature search of PubMed/Medline, EMBASE, and Google Scholar databases from 11/1/2019 till 06/07/2020. databases using following search terms or keywords: “(COVID) AND (Clinical); ((heart) OR (myocard*)) AND (COVID); (COVID) AND (Troponin); (Coronavirus) AND (Heart).”

**Study Selection:** We identified all relevant studies reporting cardiovascular comorbidities, cardiac biomarkers, disease severity, and survival in COVID-19 patients.

**Data Extraction and Synthesis:** We followed preferred reporting items for systematic reviews and meta-analyses (PRISMA) guidelines for abstracting data. Pooled data was meta-analyzed using random-effects model and between-study heterogeneity was calculated with Higgins I^2^ statistic.

**Main outcome and measures:** To assess the impact of HTN and CHF and to evaluate different cardiac biomarkers in COVID-19 patients based on their disease severity.

**Results:** We collected pooled data on 5,967 COVID-19 patients from 20 individual studies. We found that both non-survivors and those with severe disease had an increased risk of acute cardiac injury and cardiac arrhythmias, our pooled relative risk (RR) was — 8.52 (95% CI 3.63– 19.98) (p<0.001); and 3.61 (95% CI 2.03–6.43) (p=0.001), respectively. Mean difference in the levels of Troponin-I, CK-MB, and NT-proBNP was higher in deceased and severely infected patients. The RR of in-hospital mortality was 2.35 (95% CI 1.18–4.70) (p=0.022) and 1.52 (95% CI 1.12–2.05) (p=0.008) among patients who had pre-existing CHF and hypertension, respectively.

**Conclusion and Relevance:** Cardiac involvement in COVID-19 infection appears to significantly adversely impact patient prognosis and survival. Pre-existence of CHF, and high cardiac biomarkers like NT-pro BNP and CK-MB levels in COVID-19 patients correlates with worse outcomes.

## Introduction

COVID-19 is caused by the Severe Acute Respiratory Syndrome Coronavirus 2 (SARS-CoV2) virus that infects cells via membrane bound angiotensin converting enzyme-2 (ACE 2) receptors.^1^ Multiple studies have reported the spectrum of clinical manifestations of the disease and highlighted the involvement of the cardiovascular system.^1,2^ The prevalence of cardiovascular disease (CVD) in patients with COVID-19 is reported to range from 4% to 40% and the evidence is growing that its presence is linked with unfavorable outcomes, including, but not limited to ICU admissions and increased mortality.^3–7^ A recent metanalysis showed increased all-cause mortality and risk of severe form of COVID-19 infection in patients with underlying CVD.^8^ There are several small studies showing elevated biomarkers in COVID-19 patients suggesting myocardial injury. However, there are conflicting data regarding the association of cardiac biomarkers with the severity of disease.^4,9^ There is a paucity of data on outcomes in COVID-19 patients with underlying congestive heart failure (CHF).^10,11^ We conducted this study to delineate the influence of various cardiac conditions (especially CHF) on outcomes of patients with COVID-19 and to determine the prognostic value of cardiac biomarkers — Troponin I, Creatine Kinase-MB (CK-MB), and N-terminal-proB-type Natriuretic Peptide (NT-proBNP).

## Methods

This systematic review was conducted by following the guidelines laid down by preferred reporting items for systematic reviews and meta-analyses (PRISMA).^12^

### Objectives

1. To evaluate the demographics, and occurrence of cardiac injury and arrhythmias in patients with COVID-19.
2. To assess the impact of hypertension (HTN) and CHF on mortality in patients with COVID-19 infection.
3. To evaluate the levels of different cardiac biomarkers in subsets of COVID-19 patients based on their severity of illness (severe or non-severe) and final outcome (alive or deceased).

### Search strategy

We systematically searched PubMed/ Medline (https://pubmed.ncbi.nlm.nih.gov), EMBASE, and Google Scholar databases using following search terms or keywords: “(COVID) AND (Clinical); ((heart) OR (myocard*)) AND (COVID); (COVID) AND (Troponin); (Coronavirus) AND (Heart).” Two authors (S.L. and T.D.) independently reviewed 3427 citations to identify the number of studies satisfying our inclusion criteria. Of these, 702 were duplicates; 1647 were correspondence letters, case reports, and review articles; and 621 articles that were irrelevant to the study question based on their titles and abstracts, and hence, were excluded from the review. Abstracts and full-length articles of 457 studies were then evaluated and 38 of them were included for the qualitative synthesis. Of these, 17 articles did not report biomarkers of our interest leaving 21 studies to be included for the quantitative assessment (Figure 1). One study was later on excluded owing to article retraction, leaving a total of 20 studies for our final meta-analysis. Any conflicts regarding the study selection were resolved by a mutual consensus.^13^ All in-vivo studies on animals or tissue samples were excluded.

**Figure 1.**
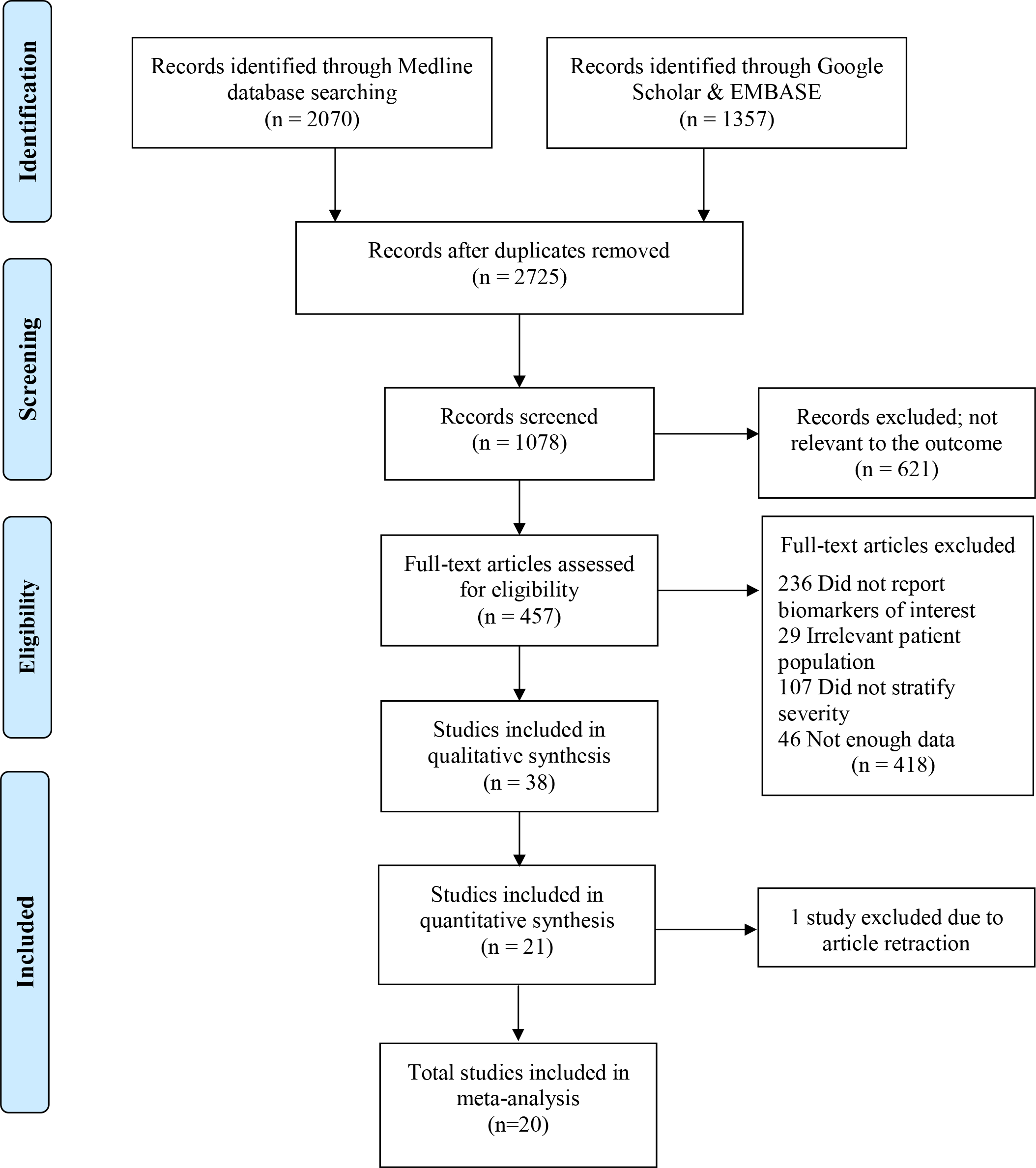
PRISMA flow diagram for the search and inclusion strategy.

### Eligibility criteria and study selection

All the published studies between December 2019 and May 2020 that reported the COVID-19 patients’ characteristics, their co-existing comorbidities, and laboratory data stratified on the basis of patient outcome and/or disease severity were included in this systematic review. Our exclusion criteria were confined to the articles that either did not report laboratory values of cardiac biomarkers or did not classify patients’ outcomes based on the severity of infection or the final patient outcome.

### Data extraction and quality assessment

Two authors (S.L. and T.D.) extracted the data. We collected the following variables: Author(s) names; year of publication; study design; duration of study period; sample population; demographic and baseline characteristics of patients; laboratory biomarkers to identify cardiac injury (namely — CK-MB, cardiac sensitive troponin I, NT-proBNP); disease severity (severe/ non-severe); and in-hospital patient outcomes (alive/deceased). The criteria used by authors for classifying patients as severe or non-severe is provided in eTable 1 in *Supplement*. Acute cardiac injury is often identified in the presence of elevated serum levels of high-sensitivity troponin^14^, however no specific cut-off value exists.

**Table 1.**
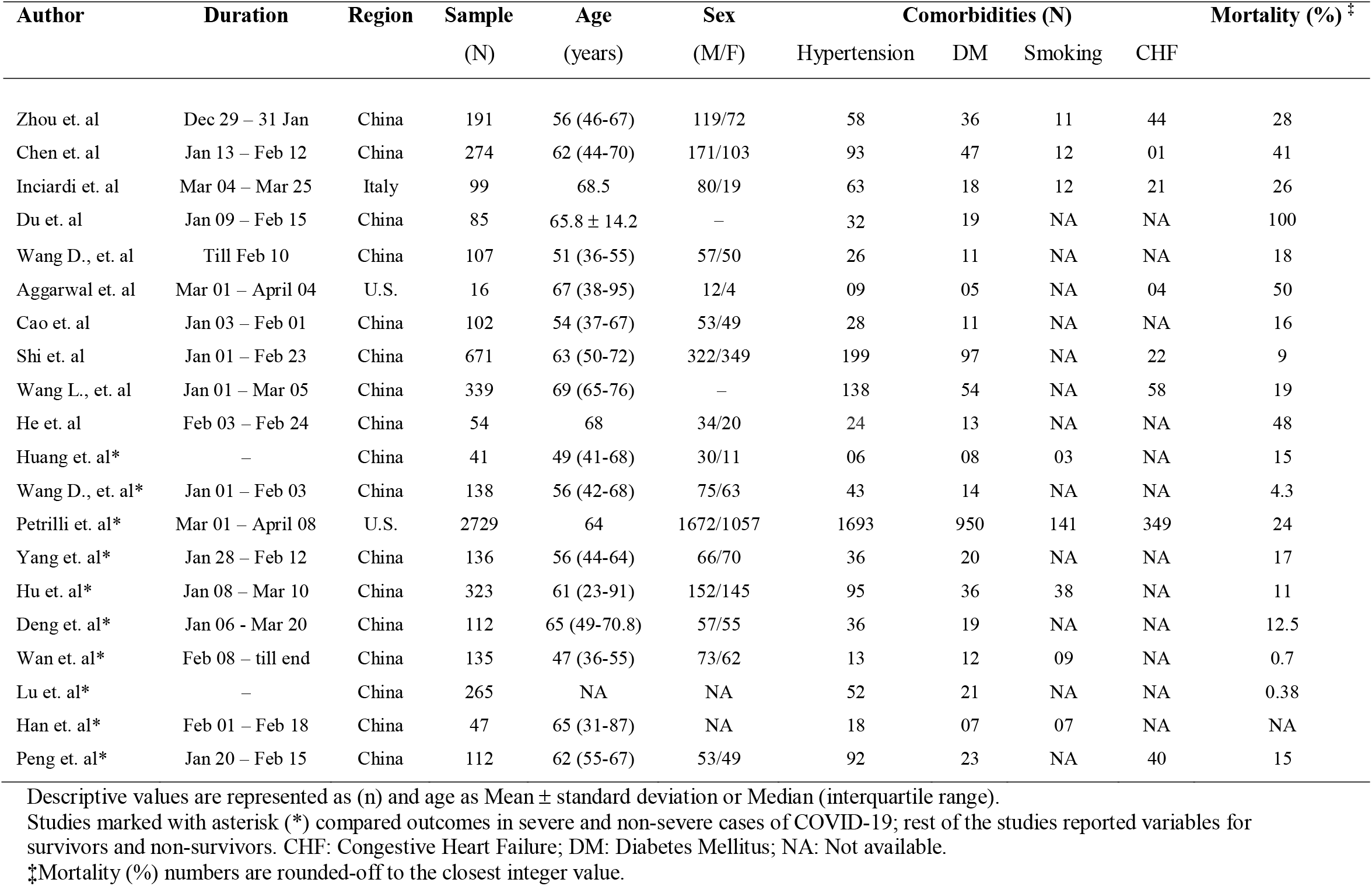
Patient demography and details of studies and in the systematic review.

| Author | Duration | Region | Sample<br>(N) | Age<br>(years) | Sex<br>(M/F) | Comorbidities (N) |  |  |  | Mortality (%) <sup>‡</sup> |
| --- | --- | --- | --- | --- | --- | --- | --- | --- | --- | --- |
|  |  |  |  |  |  | Hypertension | DM | Smoking | CHF |  |
| Zhou et. al | Dec 29 – 31 Jan | China | 191 | 56 (46-67) | 119/72 | 58 | 36 | 11 | 44 | 28 |
| Chen et. al | Jan 13 – Feb 12 | China | 274 | 62 (44-70) | 171/103 | 93 | 47 | 12 | 01 | 41 |
| Inciardi et. al | Mar 04 – Mar 25 | Italy | 99 | 68.5 | 80/19 | 63 | 18 | 12 | 21 | 26 |
| Du et. al | Jan 09 – Feb 15 | China | 85 | 65.8 ± 14.2 | – | 32 | 19 | NA | NA | 100 |
| Wang D., et. al | Till Feb 10 | China | 107 | 51 (36-55) | 57/50 | 26 | 11 | NA | NA | 18 |
| Aggarwal et. al | Mar 01 – April 04 | U.S. | 16 | 67 (38-95) | 12/4 | 09 | 05 | NA | 04 | 50 |
| Cao et. al | Jan 03 – Feb 01 | China | 102 | 54 (37-67) | 53/49 | 28 | 11 | NA | NA | 16 |
| Shi et. al | Jan 01 – Feb 23 | China | 671 | 63 (50-72) | 322/349 | 199 | 97 | NA | 22 | 9 |
| Wang L., et. al | Jan 01 – Mar 05 | China | 339 | 69 (65-76) | – | 138 | 54 | NA | 58 | 19 |
| He et. al | Feb 03 – Feb 24 | China | 54 | 68 | 34/20 | 24 | 13 | NA | NA | 48 |
| Huang et. al* | – | China | 41 | 49 (41-68) | 30/11 | 06 | 08 | 03 | NA | 15 |
| Wang D., et. al* | Jan 01 – Feb 03 | China | 138 | 56 (42-68) | 75/63 | 43 | 14 | NA | NA | 4.3 |
| Petrilli et. al* | Mar 01 – April 08 | U.S. | 2729 | 64 | 1672/1057 | 1693 | 950 | 141 | 349 | 24 |
| Yang et. al* | Jan 28 – Feb 12 | China | 136 | 56 (44-64) | 66/70 | 36 | 20 | NA | NA | 17 |
| Hu et. al* | Jan 08 – Mar 10 | China | 323 | 61 (23-91) | 152/145 | 95 | 36 | 38 | NA | 11 |
| Deng et. al* | Jan 06 - Mar 20 | China | 112 | 65 (49-70.8) | 57/55 | 36 | 19 | NA | NA | 12.5 |
| Wan et. al* | Feb 08 – till end | China | 135 | 47 (36-55) | 73/62 | 13 | 12 | 09 | NA | 0.7 |
| Lu et. al* | – | China | 265 | NA | NA | 52 | 21 | NA | NA | 0.38 |
| Han et. al* | Feb 01 – Feb 18 | China | 47 | 65 (31-87) | NA | 18 | 07 | 07 | NA | NA |
| Peng et. al* | Jan 20 – Feb 15 | China | 112 | 62 (55-67) | 53/49 | 92 | 23 | NA | 40 | 15 |

Descriptive values are represented as (n) and age as Mean ± standard deviation or Median (interquartile range).
Studies marked with asterisk (\*) compared outcomes in severe and non-severe cases of COVID-19; rest of the studies reported variables for survivors and non-survivors. CHF: Congestive Heart Failure; DM: Diabetes Mellitus; NA: Not available.
<sup>‡</sup>Mortality (%) numbers are rounded-off to the closest integer value.

### Data synthesis and analyses

We classified patients under the spectrum of “fulminant” based on severity of their disease progression including death, whereas survivors and non-severe cases were collectively classified under “non-fulminant” spectrum of illness. We also compared the relative risk of death during index admission to the hospital between patients with and without pre-existing CHF.

Categorical variables between the patient groups were summarized using the Manel-Haenszel risk ratios (RR)^15,16^ while the continuous data variables were summarized using the mean difference (MD), along with their corresponding 95% CI. These pooled estimates were calculated by using the random-effects meta-analysis model. For calculating tau-square (τ^2^), we used Hartung-Knapp-Sidik-Jonkman (HKSJ) method over the traditional DerSimonian-Laird method, as it is known to perform better with fewer number of studies and has lower type-I error rates even when combining studies with unequal sample size.^17^ Studies that had reported descriptive values as median (interquartile range) were transformed to mean ± standard deviations by using Wan method.^18^

Publication bias was evaluated by constructing funnel-plots of the effect size against its standard error (SE) and performing Egger’s linear regression test of funnel-plot asymmetry. Between-study heterogeneity was calculated with Higgins I^2^ statistic. A two-sided p value < 0.05 was considered statistical significance. We used “meta” and “metafor” packages for conducting our meta-analyses.^19,20^ Statistical analyses were conducted by S.L. in R (v3.6.3) and reviewed by S.R and Z.S.

## Results

### Baseline Characteristics

Overall, we collected 5,967 patients from 20 individual study reports. Table 1 outlines the details of studies and demographics of patients included in this systematic review. The age of patients ranged from 23–95 years, with a prevalence of females ranging from 19%–52%. The prevalence of hypertension, diabetes mellitus, and smoking was observed to be ranging from 10%–82%, 8%–24%, and 4%–14%, respectively. Ten studies reported their findings stratified according to patient outcome^6,21–29^ and 10 studies reported findings in severe and non-severe COVID-19 patients. ^3,4,9,30–36^

### Assessment of underlying cardiac conditions

We found that both non-survivors and those who had severe COVID-19 infection (fulminant group) had an 8-fold increased risk of acute cardiac injury compared to survivors/non-severe patients (non-fulminant group); pooled RR was 8.52 (95% CI 3.63–19.98) (p<0.001) (Figure 2). A similar trend of increased risk was observed for any cardiac arrhythmia; pooled RR was 3.61 (95% CI 2.03–6.43) (p=0.001) (Figure 2).

**Figure 2.**
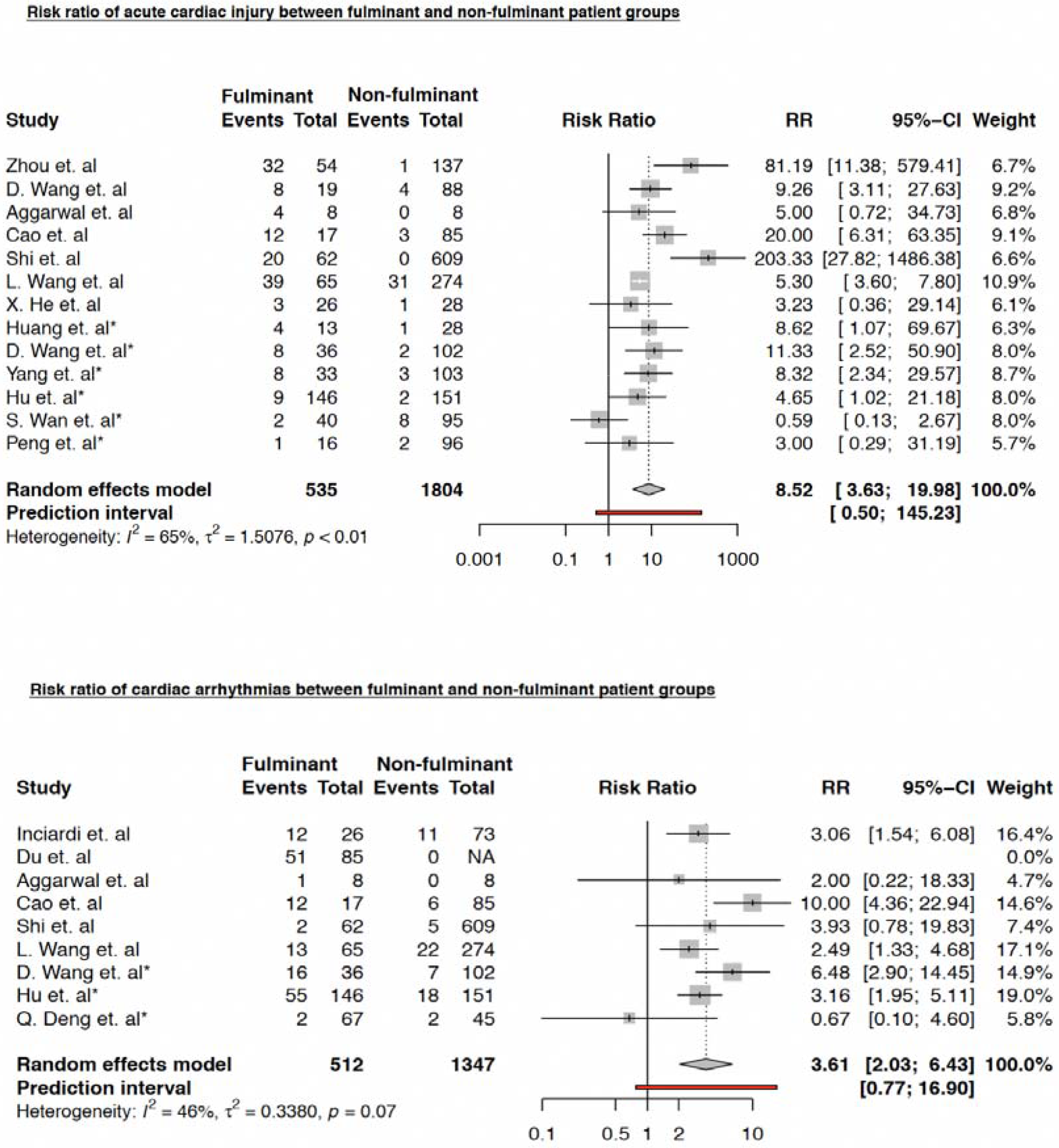
Risk ratio (RR) of acute cardiac injury (ACI) in fulminant vs non-fulminant patient groups. Studies marked with asterisk (*) compared outcomes in severe and non-severe cases of COVID-19.

We also found that the risk of death was significantly higher in CHF patients; RR 3.38 (95% CI 1.80–6.32) (p=0.004); however, the risk of in-hospital death in patients with underlying hypertension was not found to be statistically significant; RR 1.57 (95% CI 0.97–2.56) (p=0.06) (Figure 3).

**Figure 3.**
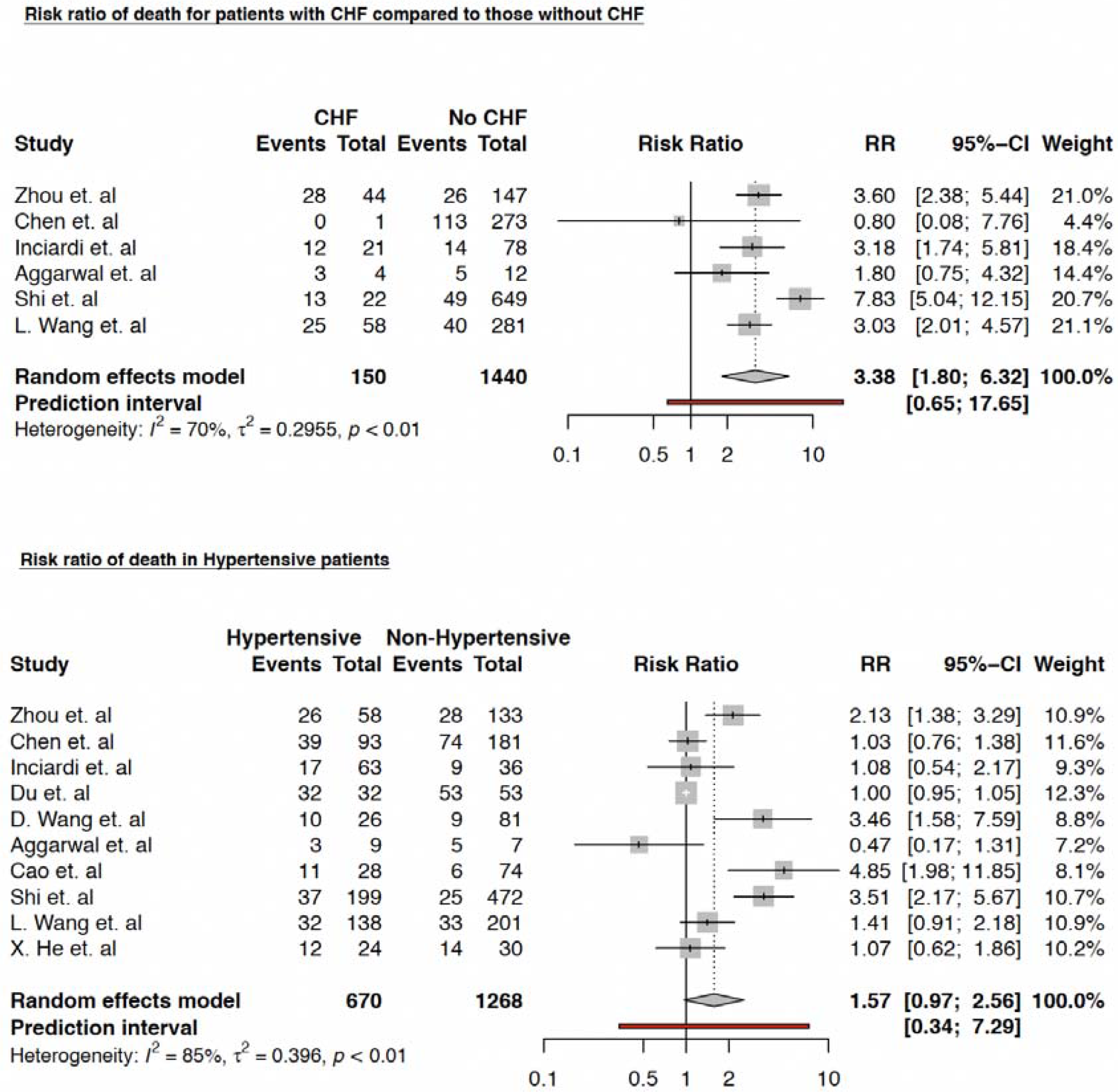
Relative risk of in-hospital death between patients — with and without pre-existing congestive heart failure; and — with and without pre-existing hypertension.

### Trends in the serum levels of cardiac biomarkers

Levels of Troponin-I were higher in fulminant group but did not reach statistical significance; mean difference (MD) was 77.93 pg/mL (95% CI -6.47–162.33) (p=0.067). The levels of CK-MB [MD 1.98 ng/mL (95% CI 0.72–3.24) (p=0.012)] and NT-proBNP [MD 1141.73 pg/mL (95% CI 336.60–1946.86) (p=0.011)] were found to be significantly higher among the patient groups with severe COVID-19 infection across individual studies. Trends of cardiac biomarkers are summarized in forest plots in Figure 4.

**Figure 4.**
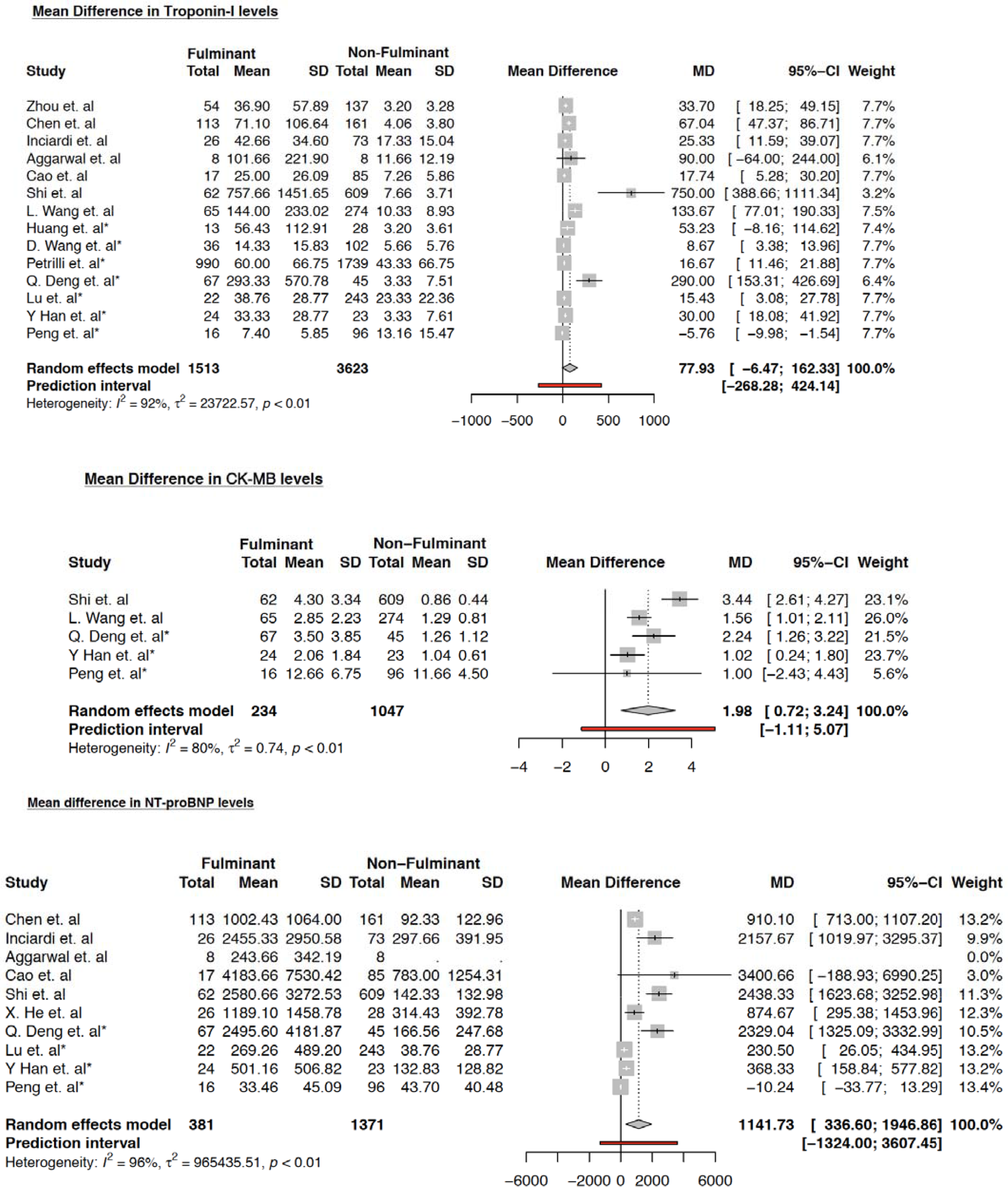
Forrest plot showing the mean differences in serum levels of cardiac biomarkers among varying patient groups. Studies marked with asterisk (*) compared severe and non-severe COVID-19 cases.

To elicit a better understanding of confounding effects of other variables, we used meta-regression models of cardiac biomarkers, adjusting for patient’s age, hypertension, diabetes, and smoking status. Our regression model showed statistically significant association in the mean difference of serum levels of NT-pro BNP only with diabetes mellitus status of the patients. (eTable 2 *in Supplement*).

## Discussion

We observed that patients who suffered from severe COVID-19 infection and non-survivors (fulminant group) had a substantially higher burden of cardiovascular diseases compared to non-severe cases and survivors (non-fulminant group). These patients were also found to be at an increased risk of acute cardiac injury and cardiac rhythm irregularities, likely owing to the involvement of myocardium in coronavirus infection which is evidenced by the elevated levels of cardiac biomarkers. Our results are consistent with other meta-analyses, which concluded that patients with underlying cardiovascular disorders have a higher propensity to develop severe and fulminant COVID-19 infection.^37,38^ Furthermore, in a latest study, Lala et. al found that the presence of cardiovascular diseases in COVID-19 patients increases their susceptibility to develop myocardial damage, and is associated with a higher mortality risk.^39^ Although, the prior systemic reviews and metanalyses have looked into the combined effect of underlying cardiovascular comorbidities on COVID-19 outcomes, this study is novel as we largely focused on impact of isolated CHF on COVID-19 outcomes. Also, prior studies had patients mostly from Southeast Asia, whereas, we incorporated patients from other geographic regions as well.

COVID-19 is a novel infectious disease caused by Severe Acute Respiratory syndrome coronavirus 2 (SARS-CoV-2) that binds to angiotensin converting enzyme 2 (ACE 2) receptors to gain entry into the human body. ^1,14^ These receptors are present on various organs including lung alveolar epithelial cells, vascular endothelial cells and cardiac myocytes and pericytes.^40–42^ The precise mechanisms for myocardial injury in COVID-19 cases are unclear at the moment. However, there are 2 proposed hypotheses — myocardial insult could either be due to “cytokine storm” suggested by elevated levels of interleukin-6, lactate dehydrogenase among a few other biomarkers; or due to the direct effect on the myocardium by causing a down-regulation of the expression of ACE2 protein expression within the cardiac myocytes. This ACE2 protein is speculated to have a protective effect against myocardial and lung injury by reducing This ACE2 protein is speculated to have a protective effect against myocardial and lung injury by reducing blood pressure, inflammation and fibrosis.^1,43–45^

Several rhythm abnormalities like atrioventricular block, atrial fibrillation, polymorphic ventricular tachycardia and pulseless electrical activity have been associated with COVID-19.^46^ The exact mechanism of underlying rhythm abnormalities is still not completely elucidated. Myocarditis has been associated with COVID-19, and elevation of inflammatory biomarkers can also suggest direct involvement of electrical conduction system.^47^ Moreover, the medications used in treatment of COVID-19 patients can increase QTc-interval and cause life threatening arrhythmias.^48^ This presence of inflammatory milieu and direct myocardial insult might have led to increased risk of cardiac arrhythmias as noted in our study.

We also observed that presence of heart failure is associated with increased mortality among COVID-19 patients. This is consistent with other studies who have shown similar findings.^21^ ACE 2 counteracts the effect of angiotensin II in states with excessive activation of renin-angiotensin system like heart failure.^1^ CHF patients are known to have dysregulation of intracellular calcium handling mechanism^49^ and COVID-19 can cause hypoxia induced excessive intracellular calcium leading to cardiac myocyte apoptosis.^50^ This can lead to increased mortality observed in patients with CHF.

NT-pro BNP has been known to be elevated in patients with CHF^51^ and its elevation is associated with poor prognosis in patients with sepsis, pneumonia.^52^ Increased ventricular wall stress due to hypoxia induced pulmonary hypertension, impaired clearance in critically ill patients with renal failure are a few mechanisms which can lead to elevation of NT-pro BNP levels in COVID-19 patients.^53^ High NT-pro BNP levels had been associated with increased mortality in COVID-19 patients.^54^ A recent systemic review which predominantly included patient population from China showed a similar finding of increased mortality.^55^ Although we had patients from different geographical regions of the world, yet these findings were consistent with our study.

## Limitations

Our systematic review inevitably has certain limitations such as between-study heterogeneity and the potential publication bias. Another concerning aspect is the retrospective study designs of included researches, and the lack of individual patient severity and medical information (such as smoking status), because of which we were unable to adjust the confounding effects of comorbidities and other variables.

## Conclusion

In summary, the presence of CHF in patients with COVID-19 is associated with increased mortality and worse outcomes during index admission. There is increased risk of acute cardiac injury and cardiac arrhythmia among non-survivors and severe COVID-19 patients. Elevated cardiac biomarkers like NT-pro BNP and CK-MB are associated with poor prognosis and can help identify “at-risk” COVID-19 patients. We suggest close monitoring of severe COVID-19 patients with underlying cardiac comorbidities to reduced mortality.

## Data Availability

N/A

## Acknowledgements

None.

